# Acute kidney injury in patients with non-valvular atrial fibrillation treated with rivaroxaban or warfarin: a population-based study from the United Kingdom

**DOI:** 10.1101/2022.04.17.22273705

**Authors:** Antonio González-Pérez, Yanina Balabanova, María E Sáez, Gunnar Brobert, Luis A Garcia Rodriguez

## Abstract

Acute kidney injury (AKI) has been associated with use of oral anticoagulants (OACs) in patients with non-valvular atrial fibrillation (NVAF). We aimed to compare AKI risk among users of rivaroxaban vs. warfarin. We identified two cohorts of NVAF patients who initiated rivaroxaban (15/20 mg/day, N=6436) or warfarin (N=7129) excluding those without estimated glomerular filtration rate values recorded in the year before OAC initiation and those with a history of end-stage renal disease or AKI. We used two methods to define AKI during follow-up (mean 2.5 years): coded entries (method A), and the Aberdeen AKI phenotyping algorithm (method B), using recorded renal function laboratory values during the study period to identify a sudden renal deterioration event. Cox regression was used to calculate hazard ratios (HRs) for AKI with rivaroxaban vs. warfarin use, adjusted for confounders. The number of identified incident AKI cases was 249 (method) A and 723 (method B). Of the latter, 104 (14.4%) were also identified by method A. After adjusting for age, sex, baseline renal function and comorbidity, HRs (95% CIs) for AKI were 1.20 (0.93–1.55; *p*=0.16) using method A, and 0.80 (0.68–0.93; *p*<0.01) using method B. Estimates stratified by baseline level of chronic kidney disease were largely consistent with the main estimates. In conclusion, our results support a beneficial effect of rivaroxaban over warfarin in terms of AKI occurrence in patients with NVAF. More research into how best to define AKI using primary care records would be valuable for future studies.

## Introduction

Individuals with atrial fibrillation (AF) have a five-fold increased risk of stroke compared with those without the condition[1] and most require long-term prophylaxis with an oral anticoagulant (OAC) to effectively mitigate this risk. Although direct oral anticoagulants (DOACs) are recommended first-line OAC therapy in these patients,[2-4] vitamin K antagonists (VKAs) remain extensively used in clinical practice.[5]

The last decade has seen emerging evidence of deterioration in renal function in patients receiving oral anticoagulants. Initially reported in patients receiving warfarin, this clinical syndrome has been called warfarin-related nephropathy (WRN). It is defined as an acute increase in international normalised ratio (INR) to above 3.0, with evidence of acute kidney injury (AKI) within a week of this increase that is not explained by other factors, but is seemingly related to anticoagulant overdosing.[6] Reports also suggest that patients with chronic kidney disease (CKD), which is prevalent in over a third of patients with AF,[7] are particularly susceptible to WRN.[6] Furthermore, WRN has been associated with accelerated CKD progression and as a significant risk factor for short- and long-term mortality.[6, 8] There have also been reports of AKI associated with DOACs,[8] although the evidence suggests that risks are greater with warfarin.[9-12] Use of large population-based longitudinal healthcare databases are suitable data sources for conducting pharmacoepidemiological research. However, AKI can be challenging to identify from many secondary data sources due to limitations related to coding and availability of reliable laboratory data. In a study of AKI incidence using electronic health record (EHR) data from the United Kingdom, Sawhney *et al*[13], found that while the majority of individuals developing AKI could be identified from hospital tests alone, around half were identified when using an alternative method of case identification based on the Kidney Disease Improving Global Outcomes phenotyping algorithm code. Additionally, several claims database studies on this topic have had limited information on some confounders. The primary objective of this study was to compare the incidence of AKI between new users of rivaroxaban and new users of warfarin in UK primary care. The secondary objective was a stratified analysis according to baseline renal function.

## Materials and methods

### Study design and data source

We undertook a population-based cohort study using anonymised longitudinal primary care electronic health records (EHRs) from the IQVIA Medical Research Data-UK (IMRD-UK). The database covers approximately 6% of the UK population,[14] is representative of the UK demographic,[15] and has been validated for pharmacoepidemiology research.[16] The data captured are those that are entered in routine clinical practice, including coded diagnoses (Read codes),[17] referrals, results of laboratory tests and all prescriptions issued. Primary care practitioners (PCPs) can also enter information manually in a free text field. Data communicated back from secondary care are entered in the patient’s primary care record retrospectively. The study protocol was approved by an independent scientific research committee (reference SRC-20SRC023). Data collection for IMRD-UK was approved by the South East Multicentre Research Ethics Committee in 2003 and individual studies using IMRD-UK data do not require separate ethical approval if only anonymised data are used.

### Source population and study cohorts

Identification of the study cohorts is shown in **Fig 1**. The source population comprised all individuals aged ≥18 years with NVAF and a first prescription (start date) for rivaroxaban (15 or 20 mg/day) or warfarin between 1 January 2014 and 31 March 2019. Individuals could only enter the source population if they were OAC naïve, and if they had been registered with their participating practice with a first prescription for any medication at least 1 year previously. Owing to the absence of a Read code specifically for NVAF, to identify patients with NVAF we identified those with a Read code for AF any time before the start date or within the 2 weeks after, excluding those with a code for heart valve replacement or mitral stenosis (i.e. valvular AF) during the same time period. For this present study, we excluded individuals without an estimated glomerular filtration rate (eGFR) value recorded in the year before the start date, and those with a history of AKI (using method A and/or B [i.e. either AKI codes or AKI based on recorded laboratory measurements of renal parameters], as described later) or end-stage renal disease (ESRD).

**Fig 1.**
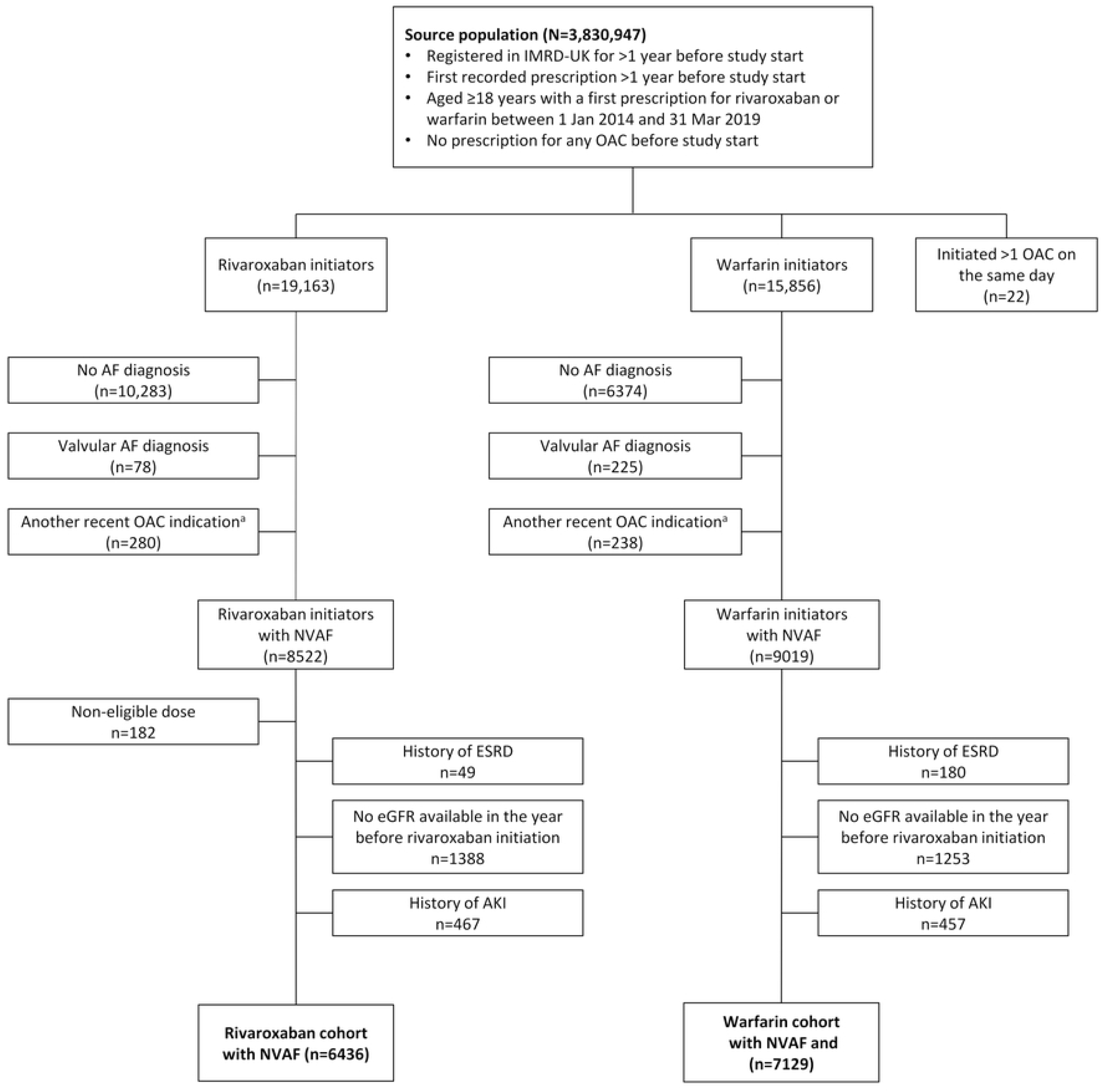
Flowchart illustrating the identification of the study cohorts. ^a^A record of VTE or orthopaedic arthroplasty in the 3 months before the first OAC prescription or in the week after. AF, atrial fibrillation; eGFR, estimated glomerular filtration rate; IMRD, IQVIA Medical Research Data; NVAF, non-valvular atrial fibrillation, OAC, oral anticoagulant; SCr, serum creatinine

### Follow-up and AKI identification

We followed the rivaroxaban and warfarin cohorts from the start date until the earliest of the following: a record of AKI, death, the last date of data collection for their practice, or the end of the study period (30 September 2019). We used two approaches to identify AKI regardless of whether it was a nosocomial or a community-acquired event. The first (method A) was based on Read code entries in the patient’s EHR indicating AKI or acute dialysis (defined as a code for dialysis and non-continuation of dialysis from 30 days after the initial dialysis code) along with a record of an outpatient visit to secondary care/hospitalisation (see **S1 Table** for codes). The second (method B) was based on recorded serum creatinine (SCr) values using the validated Aberdeen AKI phenotyping algorithm developed by Sawhney *et al*.[13] This method is potentially more accurate because it uses all recorded renal function laboratory values during the study period to identify a sudden renal deterioration event. Using method B, an AKI event was determined if any of the following three criteria were met: *criterion 1*, serum creatinine ≥1.5 times higher than the median of all creatinine values 8–365 days ago; *criterion 2*, serum creatinine ≥1.5 times higher than the lowest creatinine within 7 days; *criterion 3*, serum creatinine >26 μmol/L higher than the lowest creatinine within 48 hours). We manually reviewed the EHRs for a random sample of 100 cases to confirm the AKI event.

### Covariates

We extracted data on age, sex, and Townsend index score of deprivation,[15] comorbidities and frailty (using an index developed for research using primary care databases[18]) on, or any time before, the start date. Comedications were determined from prescription data on/in the year before the start date, and polypharmacy was ascertained as the number of different medications prescribed in the month before (but not including) the start date. Healthcare use (number of primary care visits, referrals and hospitalisations) and lifestyle variables (body mass index, smoking status and alcohol consumption; using the most recent value/status) were ascertained in the year before the start date. To determine renal function at baseline, we used recorded eGFR values, expressed as mL/min/1.73m^2^, and used the most recent valid SCr value recorded in the year before the start date to apply the Chronic Kidney Disease Epidemiology (CKD-EPI) Collaboration equation,[19] with the omission of ethnicity as this is not routinely recorded. We also used coded clinical entries indicating CKD stage, and acute/chronic dialysis.

### Statistical analysis

Baseline characteristics were described using counts and percentages for categorical variables and mean with standard variation for continuous variables. Incidence rates of AKI, for both case definitions, were calculated using the number of incident cases during follow-up as the numerator and total person-years as the denominator, with 95% confidence intervals (CIs) assuming a Poisson distribution. Incidence rates were stratified by baseline renal function (>50 and ≤50 mL/min/1.73m^2^). Cox proportional hazards regression was used to calculate hazard ratios (HRs) for AKI with rivaroxaban vs. warfarin use, adjusted for confounders, both overall and stratified by baseline renal function. We performed several sensitivity analyses. Firstly, we censored patients at the date of OAC discontinuation (>30 days after the end of the last consecutive prescription of the starting OAC; on-treatment analysis). Secondly, patients were eligible to contribute person-time to different OAC exposure categories according to their current exposure irrespective of the starting drug (as-treated analysis). Thirdly, we restricted the analysis to cases of severe AKI (defined as stage 2 or 3 AKI; this was only performed for Method B because there are no Read codes that specify AKI severity). Fourthly, repeated the ITT analysis adjusting for death as a competing risk using Fine and Gray models.[20] Lastly, we used two more stringent definitions of the Aberdeen algorithm to identify AKI cases: i) meeting criterion 2 or 3, and ii) meeting criterion 3.

## Results

Baseline characteristics of the study cohorts are shown in **Table 1**. Mean age was 74 years in both cohorts, and over half were male (57% rivaroxaban cohort, 56% warfarin cohort). Individuals in the warfarin cohort more likely to have a higher level of deprivation and history of ischaemic heart disease, while individuals in the rivaroxaban cohort were more likely to be severely frail and have a higher number of recent referrals/hospitalisations (in the previous year).

**Table 1.** Baseline characteristics of the study cohorts.

|  | **Rivaroxaban cohort**<br>**N=6436** | **Warfarin cohort**<br>**N=7129** |
| --- | --- | --- |
| **Age (years)** |  |  |
| 18–49 | 108 (1.7) | 101 (1.4) |
| 50–59 | 458 (7.1) | 391 (5.5) |
| 60–69 | 1349 (21.0) | 1574 (22.1) |
| 70–79 | 2357 (36.6) | 2857 (40.1) |
| 80–89 | 1763 (27.4) | 1972 (27.7) |
| ≥90 | 401 (6.2) | 234 (3.3) |
| Mean (SD) | 74.4 (10.6) | 74.2 (9.6) |
| **Sex** |  |  |
| Male | 3687 (57.3) | 4011 (56.3) |
| Female | 2749 (42.7) | 3118 (43.7) |
| **Townsend index** |  |  |
| 1st quintile (least deprived) | 1288 (20.0) | 1395 (19.6) |
| 2nd quintile | 1290 (20.0) | 1527 (21.4) |
| 3rd quintile | 1195 (18.6) | 1220 (17.1) |
| 4th quintile | 896 (13.9) | 1116 (15.7) |
| 5th quintile (most deprived) | 510 (7.9) | 674 (9.5) |
| Missing | 1257 (19.5) | 1197 (16.8) |
| **Polypharmacy** |  |  |
| 0–4 | 1136 (17.7) | 1081 (15.2) |
| 5–9 | 3493 (54.3) | 3979 (55.8) |
| ≥10 | 1807 (28.1) | 2069 (29.0) |
| **Smoking** |  |  |
| Non-smoker | 2693 (41.8) | 2818 (39.5) |
| Smoker | 503 (7.8) | 592 (8.3) |
| Ex-smoker | 3233 (50.2) | 3718 (52.2) |
| Missing | 7 (0.1) | 1 (0.0) |
| **BMI** |  |  |
| <20 | 202 (3.1) | 181 (2.5) |
| 20–24 | 1358 (21.1) | 1426 (20.0) |
| 25–29 | 2283 (35.5) | 2532 (35.5) |
| ≥30 | 2335 (36.3) | 2758 (38.7) |
| Missing | 258 (4.0) | 232 (3.3) |
| Mean (SD) | 29.1 (6.2) | 29.5 (6.4) |
| **Comorbidities/previous clinical history** |  |  |
| IHD | 1524 (23.7) | 1866 (26.2) |
| Cancer | 1018 (15.8) | 1131 (15.9) |
| Diabetes | 1375 (21.4) | 1601 (22.5) |
| Heart failure | 720 (11.2) | 901 (12.6) |
| AKI (history) | 0 (0.0) | 0 (0.0) |
| **eGFR at baseline (mL/min/1.73m<sup>2</sup>)** |  |  |
| >50 | 5547 (86.2) | 6043 (84.8) |
| 30–50 | 829 (12.9) | 961 (13.5) |
| <30 | 60 (0.9) | 125 (1.8) |
| Mean (SD) | 70.5 (52.5) | 70.4 (78.8) |
| **eGFR at baseline**<br>**(measurements)** |  |  |
| 1 | 128 (2.0) | 149 (2.1) |
| 2–5 | 918 (14.3) | 1039 (14.6) |
| 6–11 | 1654 (25.7) | 1791 (25.1) |
| 12–23 | 2403 (37.3) | 2728 (38.3) |
| ≥24 | 1333 (20.7) | 1422 (19.9) |
| Mean (SD) | 16.6 (14.8) | 16.5 (14.6) |
| **eGFR follow-up**<br>**(measurements)** |  |  |
| None | 762 (11.8) | 746 (10.5) |
| 1 | 711 (11.0) | 549 (7.7) |
| 2–5 | 2470 (38.4) | 2295 (32.2) |
| 6–11 | 1579 (24.5) | 1931 (27.1) |
| 12–23 | 697 (10.8) | 1159 (16.3) |
| ≥24 | 217 (3.4) | 449 (6.3) |
| Mean (SD) | 6.1 (7.4) | 8.4 (11.1) |
| **CHA<sub>2</sub>DS<sub>2</sub>VASc** |  |  |
| 0–1 | 903 (14.0) | 829 (11.6) |
| 2 | 1229 (19.1) | 1358 (19.0) |
| 3 | 1546 (24.0) | 1806 (25.3) |
| 4 | 1450 (22.5) | 1658 (23.3) |
| 5 | 800 (12.4) | 900 (12.6) |
| ≥6 | 508 (7.9) | 578 (8.1) |
| Mean (SD) | 3.2 (1.6) | 3.3 (1.6) |
| **Frailty** |  |  |
| Fit | 1095 (17.0) | 1136 (15.9) |
| Mild frailty | 2672 (41.5) | 3081 (43.2) |
| Moderate frailty | 1871 (29.1) | 2138 (30.0) |
| Severe frailty | 798 (12.4) | 774 (10.9) |
| **PCP visits** |  |  |
| 0–4 | 50 (0.8) | 43 (0.6) |
| 5–9 | 587 (9.1) | 602 (8.4) |
| 10–19 | 2720 (42.3) | 3051 (42.8) |
| ≥20 | 3079 (47.8) | 3433 (48.2) |
| **Referrals** |  |  |
| 0–9 | 3824 (59.4) | 4434 (62.2) |
| 10–19 | 2064 (32.1) | 2174 (30.5) |
| ≥20 | 548 (8.5) | 521 (7.3) |
| **Hospitalisations** |  |  |
| 0 | 3573 (55.5) | 4236 (59.4) |
| 1 | 1471 (22.9) | 1493 (20.9) |
| ≥2 | 1392 (21.6) | 1400 (19.6) |

Data are n (%) unless otherwise stated.
AKI, acute kidney injury; BMI, body mass index; eGFR, estimated glomerular filtration rate; IHD, ischaemic heart disease; PCP, primary care practitioner; SD, standard deviation

After a mean follow-up of 2.5 years, the number of incident AKI cases identified using coded entries (Method A) was 136 (rivaroxaban cohort) and 113 (warfarin cohort), and using the Aberdeen algorithm (Method B) was 266 (rivaroxaban cohort) and 457 (warfarin cohort). Of these 723 AKI cases identified by the Aberdeen algorithm, 104 (14.4%) were also identified by codes (**S1 Fig**); of the 249 AKI cases identified by codes, 145 (58.2%) were not identified using the Aberdeen algorithm. Incidence rates of AKI using cases identified from coded entries (Method A) were 81.2 per 10,000 person-years (95% CI: 66.9–97.6, rivaroxaban cohort) and 67.9 per 10,000 person-years (95% CI: 57.0–80.3, warfarin cohort), and using the Aberdeen algorithm (Method B) they were 194.4 per 10,000 person-years (95% CI: 171.7–219.2) and 234.5 per 10,000 person-years (95% CI: 213.5–257.0), respectively.

In the Cox regression analysis, after adjusting for confounders, we found no clear evidence for a difference in AKI risk between rivaroxaban and warfarin users when using coded entries to identify AKI (HR 1.19, 95% CI: 0.92–1.54; p=0.18), but strong evidence that rivaroxaban users had a reduced risk of AKI vs. warfarin users when using the Aberdeen algorithm (HR 0.80, 95% CI: 0.68–0.93; p<0.01) (**Table 2**). Estimates stratified by baseline CKD (**Table 3**), and results of the sensitivity analyses were largely consistent with the overall estimates (**Supplementary S2-S8 Tables**).

**Table 2.**
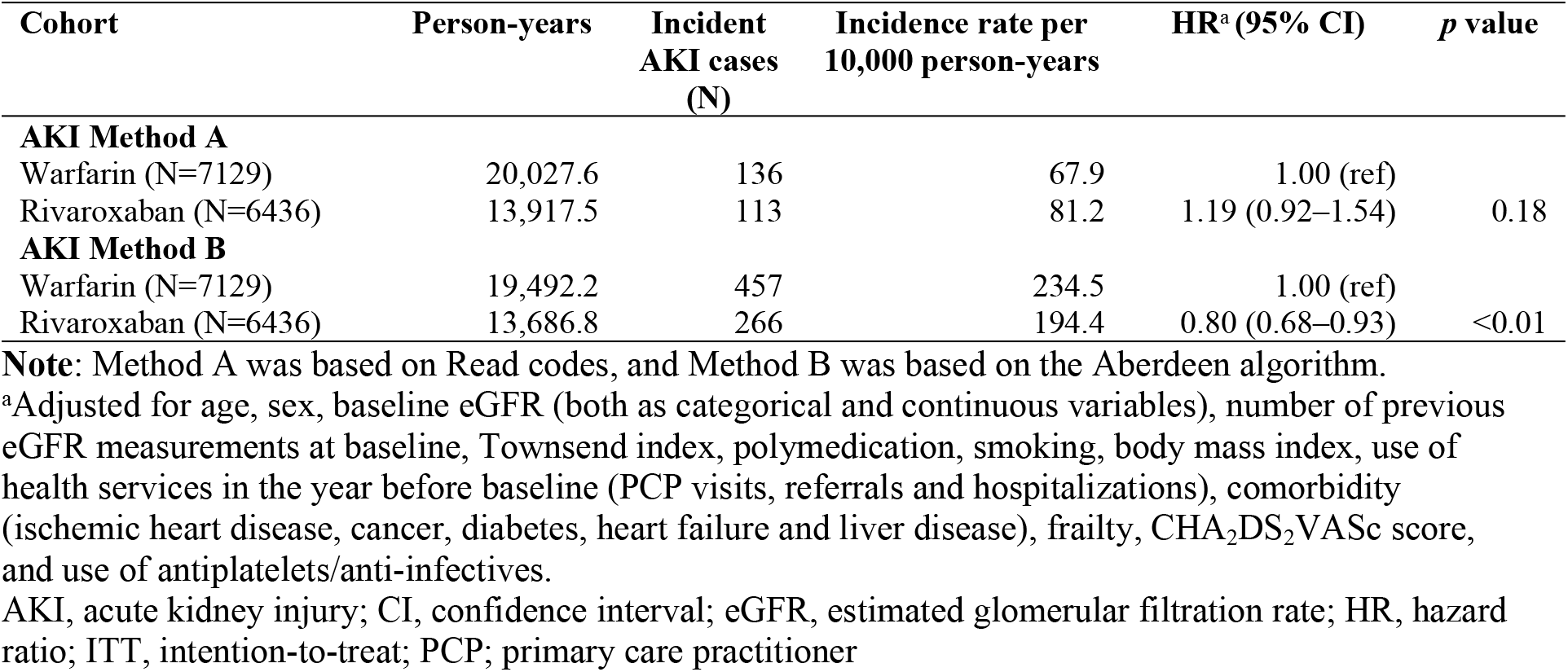
Incidence rate per 10,000 person-years of AKI, and HRs (95% CIs) comparing AKI in the rivaroxaban vs. warfarin cohorts (ITT analysis).

**Table 3.** Incidence rate per 10,000 person-years of AKI and HR (95% CI) comparing AKI in the rivaroxaban vs. warfarin cohorts, stratified by baseline renal function (ITT analysis).

| Baseline eGFR | Cohort | Person-years | Incident AKI cases (N) | Incidence rate per 10,000 person-years | HR<sup>a</sup> (95% CI) | *p* value |
| --- | --- | --- | --- | --- | --- | --- |
| AKI Method A |  |  |  |  |  |  |
| eGFR >50 mL/min/1.73m<sup>2</sup> | Warfarin (N=6043) | 17366.9 | 93 | 53.55 | 1.00 (ref) | 0.48 |
|  | Rivaroxaban (N=5547) | 12155.1 | 74 | 60.88 | 1.12 (0.82–1.53) |  |
| eGFR ≤50 mL/min/1.73m<sup>2</sup> | Warfarin (N=1086) | 2660.7 | 43 | 161.61 | 1.00 (ref) | 0.21 |
|  | Rivaroxaban (N=889) | 1762.4 | 39 | 221.29 | 1.34 (0.85–2.12) |  |
| AKI Method B |  |  |  |  |  |  |
| eGFR >50 mL/min/1.73m<sup>2</sup> | Warfarin (N=6043) | 16946.4 | 349 | 205.94 | 1.00 (ref) | 0.01 |
|  | Rivaroxaban (N=5547) | 11963.1 | 208 | 173.87 | 0.79 (0.66–0.94) |  |
| eGFR ≤50 mL/min/1.73m<sup>2</sup> | Warfarin (N=1086) | 2545.8 | 108 | 424.23 | 1.00 (ref) | 0.23 |
|  | Rivaroxaban (N=889) | 1723.6 | 58 | 336.5 | 0.82 (0.59–1.14) |  |

**Note:** Method A was based on Read codes, and Method B was based on the Aberdeen algorithm.
<sup>a</sup>Adjusted for age, sex, baseline eGFR (both as categorical and continuous variables), number of previous eGFR measurements at baseline, Townsend index, polymedication, smoking, body mass index, use of
health services in the year before baseline (PCP visits, referrals and hospitalizations), comorbidity (ischemic heart disease, cancer, diabetes, heart failure and liver disease), frailty, CHA<sub>2</sub>DS<sub>2</sub>VASc score, and use of antiplatelets/anti-infectives.
AKI, acute kidney injury; CI, confidence interval; eGFR, estimated glomerular filtration rate; HR, hazard ratio; ITT, intention-to-treat; PCP; primary care practitioner

## Discussion

In this large population-based EHR study of patients with NVAF, we found clear evidence of a 20% reduced risk of AKI among users of rivaroxaban compared with users of warfarin when using a definition of AKI that involved recorded laboratory renal function values. This finding was not seen when using an AKI definition based on coded database entries; however, as laboratory values are regarded as critical in AKI diagnosis, the results using the Aberdeen algorithm are likely to be more accurate. Only 14.4% of the AKI cases identified by the Aberdeen algorithm were also identified by coded AKI entries, while only 58.2% of AKI cases identified by codes were also identified by the Aberdeen algorithm, revealing poor agreement between the two case ascertainment methods.

Our findings, using the Aberdeen algorithm case ascertainment method, are consistent with several previous studies on this topic, among patients with NVAF using rivaroxaban compared with those using warfarin[9, 10, 12, 21] or phenprocoumon,[11] including among high risk groups such those with CKD[11, 12, 22], diabetes,[9, 12], heart failure[9] or the elderly.[9, 10, 21] Some have also shown similar effect sizes (15–19% reduced AKI risk), although some have reported a larger, ∼30% lower risk with rivaroxaban[9, 22] vs. warfarin, including with an even larger ∼50% lower risk seen in one study.[22] Furthermore, a recent large meta-analysis of data from randomised controlled trials (RCTs) and observational studies reported a 30% reduced risk of AKI among users of DOACs vs. warfarin.[23] Consistent with most other studies,[9, 12, 21, 23] our results did not infer any materially different effect between patients with baseline CKD and those without CKD. It is noteworthy that the AKI incidence rates in our study were much lower than those reported by Yao *et al* in their United Stated claims database study where AKI was based on primary and secondary diagnoses of AKI in linked hospitalisation data. However, as IMRD UK does include data communicated back from secondary care, we believe only a small number of cases would potentially not have been captured in the database. Our analysis focused on rivaroxaban vs. warfarin only; however, some other studies,[9, 11] albeit not all,[21-23] which have evaluated different DOACs with VKAs have reported significantly reduced risks of AKI among rivaroxaban or dabigatran users that was not seen for apixaban users. In this present study, we have also demonstrated the importance of an accurate case definition for capturing AKI events shown by the divergent results produced by the two case definitions. This also highlights the potential limitations of some databases when relying on coded entries only for clinical events that are largely defined by laboratory parameters in clinical practice.

A differential effect on AKI between rivaroxaban and warfarin is plausible due to their different mechanisms of action although the exact mechanistic pathway(s) are unclear. Warfarin affects all vitamin K dependent proteins, not just those in anticoagulation, and it has been shown to increase medial and intimal vascular calcification.^[24]^ Also, it is possible that rivaroxaban – a direct factor Xa inhibitor – could help renopreservation through a reduction in protease-activated receptor-mediated inflammation.^[24]^ Other possible pathophysiological pathways for WRN are associated with reductions in activated protein C and endothelial protein C receptor signalling.[8] Direct oral anticoagulants are excreted by the kidneys to different degrees (80% for dabigatran, 65% for rivaroxaban and 25% for apixaban), whereas VKAs are largely metabolised through the liver,[24] with relatively more harmful renal affects only being reported in the last decade. The significantly reduced AKI risk seen among rivaroxaban vs. warfarin users in our study and previous investigations is clinically meaningful considering the life-long nature of anticoagulation in patients with NVAF and the ageing population. Patients with NVAF should be managed in a way that not only effectively reduces their risk of stroke while minimising bleeding, but also best preserves renal function.

Strengths of the study include the large sample of patients with NVAF from a database representative of the UK general population, thereby enabling the calculation of precise estimates of effect and meaning our findings our generalisable to the UK as a whole. We were able to control for a wide range of potential confounders including comorbidities, co-medications, demographics and lifestyle factors, although we acknowledge the possibility of residual confounding. Furthermore, sensitivity analyses showed the results of the main analyses to be robust. A limitation of our study is the potential for some non-differential outcome misclassification due to unrecorded/incorrect laboratory measurements, biasing the risk estimates towards the null. A detection bias in the opposite direction could operate if patients on warfarin were subject to greater monitoring including more laboratory investigations, thereby increasing the likelihood of AKI diagnosis. Additionally, we did not investigate INR levels among warfarin users and were therefore unable to account for potential overdosing that could lead to renal damage in these patients, which could have led to residual confounding. Also, medication use was based on prescriptions issued that did not capture over-the-counter drug use or lack of compliance.

In conclusion, our results build on the existing evidence to support a beneficial effect of rivaroxaban over warfarin in terms of AKI occurrence in patients with NVAF. Clinicians should consider effects on renal function when making individualised decisions on the most appropriate OAC for their patients. Further evidence to support a causal association from RCTs and well-designed observational studies in other settings would help prescribers make more informed benefit–risk decisions regarding choice of long-term OAC therapy for their patients. More research into how best to define AKI using primary care EHRs would also be of value for future studies.

## Data Availability

The data underlying the results presented in the study are available from A Gonzalez-Perez (the corresponding author)

## Acknowledgments

We thank Susan Bromley. EpiMed Communications, Abingdon, UK, for medical writing assistance, funded by Bayer AG and in accordance with Good Publication Practice. IQVIA provided the IMRD-UK incorporating data from THIN, a Cegedim Database. Reference made to THIN is intended to be descriptive of the data asset licensed by IQVIA.

## Supporting information

**S1 Table**. Read codes for AKI.

**S2 Table**. Incidence rate per 10,000 person-years of AKI and HR (95% CI) comparing AKI in the rivaroxaban vs. warfarin cohorts, overall and stratified by baseline renal function: <u>on-treatment analysis.</u>

**S3 Table**. Incidence rate per 10,000 person-years of AKI and HR (95% CI) comparing AKI in the rivaroxaban vs. warfarin cohorts, overall and stratified by baseline renal function: <u>as-treated analysis.</u>

**S4 Table**. Incidence rate per 10,000 person-years of <u>severe AKI (using AKI Method B – see footnote)</u> and HR (95% CI) comparing AKI in the rivaroxaban vs. warfarin cohorts, overall and stratified by baseline renal function (intention-to-treat analysis).

**S5 Table**. Incidence rate per 10,000 person-years of <u>severe AKI (using AKI Method B – see footnote)</u> and HR (95% CI) comparing AKI in the rivaroxaban vs. warfarin cohorts, overall and stratified by baseline renal function (on-treatment analysis).

**S6 Table**. Incidence rate per 10,000 person-years of <u>severe AKI (using AKI Method B – see footnote)</u> and HR (95% CI) comparing AKI in the rivaroxaban vs. warfarin cohorts, overall and stratified by baseline renal function (as-treated analysis).

**S7 Table**. Incidence rate per 10,000 person-years of AKI and HR (95% CI) comparing AKI in the rivaroxaban vs. warfarin cohorts, stratified by baseline renal function (ITT analysis): <u>Fine and Gray models</u>.

**S8 Table**. Incidence rate per 10,000 person-years of AKI and HR (95% CI) comparing AKI in the rivaroxaban vs. warfarin cohorts, overall and stratified by baseline renal function: <u>more stringent method B criteria.</u>

**S1 Fig**. AKI cases identified by the two case identification definitions.

AKI, acute kidney injury

