## Supplementary files for "Acute kidney injury in patients with non-valvular atrial fibrillation treated with rivaroxaban or warfarin: a population-based study from the United Kingdom"

**Supplementary Table 1**. Read codes for AKI.

| **Read code** | **Description** |
|---|---|
| K04..00 | Acute renal failure |
| K040.00 | Acute renal tubular necrosis |
| K041.00 | Acute renal cortical necrosis |
| K042.00 | Acute renal medullary necrosis |
| K043.00 | Acute drug-induced renal failure |
| K044.00 | Acute renal fail urin obstruct |
| K04y.00 | Other acute renal failure |
| K04z.00 | Acute renal failure NOS |
| Kyu2000 | [X]Other acute renal failure |
| G500400 | Acute pericarditis - uraemic |
| K101000 | Acute pyelonephritis without medullary necrosis |
| K101100 | Acute pyelonephritis with medullary necrosis |
| K04..11 | ARF - Acute renal failure |
| K04..12 | Acute kidney injury |
| K043000 | Acute renal failure due to ACE inhibitor |
| K043100 | Acute renal failure induced by aminoglycoside |
| K043300 | Acute renal failure induced by cyclosporin A |
| K043400 | Acute renal failure induced by non-steroid anti-inflamm drug |
| K045.00 | Acute renal failure due to non-traumatic rhabdomyolysis |
| K046.00 | Acute renal failure induced by toxin |
| K04B.00 | Acute renal failure due to traumatic rhabdomyolysis |
| K04C.00 | Acute kidney injury stage 1 |
| K04D.00 | Acute kidney injury stage 2 |
| K04E.00 | Acute kidney injury stage 3 |

**Supplementary Table 2**. Incidence rate per 10,000 person-years of AKI and HR (95% CI) comparing AKI in the rivaroxaban vs. warfarin cohorts, overall and stratified by baseline renal function: on-treatment analysis.

| Baseline eGFR | Cohort | Person-years | Incident AKI cases (N) | Incidence rate per 10,000 person-years | HR* (95% CI) | *p* value |
|---|---|---|---|---|---|---|
| **AKI Method A** | | | | | | |
| Any eGFR | Warfarin (N=7129) | 11,201.1 | 75 | 67.0 | 1.0 (ref) | |
| | Rivaroxaban (N=6436) | 9019.2 | 70 | 77.6 | 1.18 (0.85–1.65) | 0.33 |
| eGFR >50mL/min/1.73m$^2$ | Warfarin (N=6043) | 9621.0 | 48 | 49.9 | 1.0 (ref) | |
| | Rivaroxaban (N=5547) | 7847.6 | 46 | 58.6 | 1.16 (0.77–1.76) | 0.48 |
| eGFR ≤50 mL/min/1.73m$^2$ | Warfarin (N=1086) | 1580.1 | 27 | 170.9 | 1.0 (ref) | |
| | Rivaroxaban (N=889) | 1171.7 | 24 | 204.8 | 1.15 (0.64–2.06) | 0.64 |
| **AKI Method B** | | | | | | |
| Any eGFR | Warfarin (N=7129) | 11,022.7 | 278 | 252.2 | 1.0 (ref) | |
| | Rivaroxaban (N=6436) | 8918.1 | 175 | 196.2 | 0.76 (0.62–0.92) | <0.01 |
| eGFR >50mL/min/1.73m$^2$ | Warfarin (N=6043) | 9484.6 | 206 | 217.2 | 1.0 (ref) | |
| | Rivaroxaban (N=5547) | 7760.2 | 133 | 171.4 | 0.74 (0.59–0.92) | 0.01 |
| eGFR ≤50 mL/min/1.73m$^2$ | Warfarin (N=1086) | 1538.1 | 72 | 468.1 | 1.0 (ref) | |
| | Rivaroxaban (N=889) | 1157.9 | 42 | 362.7 | 0.81 (0.54–1.20) | 0.28 |

**Note**: Method A was based on Read codes, and Method B was based on the Aberdeen algorithm.
*Adjusted for age, sex, baseline eGFR (both as categorical and continuous variables), number of previous eGFR measurements at baseline, Townsend index, polymedication, smoking, body mass index, use of health services in the year before baseline (PCP visits, referrals and hospitalizations), comorbidity (ischemic heart disease, cancer, diabetes, heart failure and liver disease), frailty, CHA2DS2VASc score, and use of antiplatelets/anti-infectives.
AKI, acute kidney injury; CI, confidence interval; eGFR, estimated glomerular filtration rate; HR, hazard ratio; PCP; primary care practitioner

**Supplementary Table 3**. Incidence rate per 10,000 person-years of AKI and HR (95% CI) comparing AKI in the rivaroxaban vs. warfarin cohorts, overall and stratified by baseline renal function: as-treated analysis.

| Baseline eGFR | Cohort | Person-years | Incident AKI cases (N) | Incidence rate per 10,000 person-years | HR* (95% CI) | *p* value |
|---|---|---|---|---|---|---|
| **AKI Method A** | | | | | | |
| Any eGFR | Warfarin (N=7230) | 14903.4 | 104 | 69.8 | 1.0 (ref) | |
| | Rivaroxaban (N=7045) | 12,064.0 | 93 | 77.1 | 1.13 (0.85–1.50) | 0.4 |
| eGFR >50 mL/min/1.73m$^2$ | Warfarin (N=6124) | 12,806.7 | 67 | 52.32 | 1.0 (ref) | |
| | Rivaroxaban (N=6081) | 10,567.3 | 62 | 58.67 | 1.12 (0.79–1.58) | 0.54 |
| eGFR ≤50 mL/min/1.73m$^2$ | Warfarin (N=1106) | 2096.7 | 37 | 176.5 | 1.0 (ref) | |
| | Rivaroxaban (N=964) | 1496.7 | 31 | 207.1 | 1.14 (0.69–1.87) | 0.61 |
| **AKI Method B** | | | | | | |
| Any eGFR | Warfarin (N=7225) | 14,580.6 | 345 | 236.6 | 1.0 (ref) | |
| | Rivaroxaban (N=7020) | 11,861.3 | 216 | 182.1 | 0.77 (0.65–0.91) | <0.01 |
| eGFR >50 mL/min/1.73m$^2$ | Warfarin (N=6119) | 12,563.2 | 259 | 206.2 | 1.0 (ref) | |
| | Rivaroxaban (N=6057) | 10,390.5 | 167 | 160.7 | 0.76 (0.63–0.93) | 0.01 |
| eGFR ≤50 mL/min/1.73m$^2$ | Warfarin (N=1106) | 2017.5 | 86 | 426.3 | 1.0 (ref) | |
| | Rivaroxaban (N=963) | 1470.9 | 49 | 333.1 | 0.82 (0.57–1.18) | 0.28 |

**Note**: Method A was based on Read codes, and Method B was based on the Aberdeen algorithm.

*Adjusted for age, sex, baseline eGFR (both as categorical and continuous variables), number of previous eGFR measurements at baseline, Townsend index, polymedication, smoking, body mass index, use of health services in the year before baseline (PCP visits, referrals and hospitalizations), comorbidity (ischemic heart disease, cancer, diabetes, heart failure and liver disease), frailty, $CHA_2DS_2VASc$ score, and use of antiplatelets/anti-infectives.

AKI, acute kidney injury; CI, confidence interval; eGFR, estimated glomerular filtration rate; HR, hazard ratio; PCP; primary care practitioner

**Supplementary Table 4**. Incidence rate per 10,000 person-years of severe AKI (using AKI Method B – see footnote) and HR (95% CI) comparing AKI in the rivaroxaban vs. warfarin cohorts, overall and stratified by baseline renal function (intention-to-treat analysis).

| **Baseline eGFR** | **Cohort** | **Person-years** | **Incident severe AKI cases (N)** | **Incidence rate per 10,000 person-years** | **HR* (95% CI)** | ***p* value** |
|---|---|---|---|---|---|---|
| Any eGFR | Warfarin (N=7129) | 19,492.2 | 104 | 53.4 | 1.0 (ref) | |
| | Rivaroxaban (N=6436) | 13,686.8 | 54 | 39.5 | 0.74 (0.53–1.04) | 0.08 |
| eGFR >50mL/min/1.73m$^2$ | Warfarin (N=6043) | 16,946.4 | 78 | 46.0 | 1.0 (ref) | |
| | Rivaroxaban (N=5547) | 11,963.1 | 38 | 31.8 | 0.68 (0.46–1.01) | 0.06 |
| eGFR ≤50 mL/min/1.73m$^2$ | Warfarin (N=1086) | 2545.8 | 26 | 102.1 | 1.0 (ref) | |
| | Rivaroxaban (N=889) | 1723.6 | 16 | 92.8 | 0.95 (0.50–1.82) | 0.88 |

**Note**: Method B was based on the Aberdeen algorithm. Severe AKI was defined as stage 2 or 3 AKI.

*Adjusted for age, sex, baseline eGFR (both as categorical and continuous variables), number of previous eGFR measurements at baseline, Townsend index, polymedication, smoking, body mass index, use of health services in the year before baseline (PCP visits, referrals and hospitalizations), comorbidity (ischemic heart disease, cancer, diabetes, heart failure and liver disease), frailty, $CHA_2DS_2VASc$ score, and use of antiplatelets/anti-infectives.

AKI, acute kidney injury; CI, confidence interval; eGFR, estimated glomerular filtration rate; HR, hazard ratio; PCP; primary care practitioner

**Supplementary Table 5**. Incidence rate per 10,000 person-years of severe AKI (using AKI Method B – see footnote) and HR (95% CI) comparing AKI in the rivaroxaban vs. warfarin cohorts, overall and stratified by baseline renal function (on-treatment analysis).

| Baseline eGFR | Cohort | Person-years | Incident severe AKI cases (N) | Incidence rate per 10,000 person-years | HR* (95% CI) | *p* value |
|---|---|---|---|---|---|---|
| Any eGFR | Warfarin (N=7129) | 11,022.7 | 63 | 57.2 | 1.0 (ref) | |
| | Rivaroxaban (N=6436) | 8918.1 | 40 | 44.9 | 0.81 (0.54–1.21) | 0.30 |
| eGFR >50mL/min/1.73m$^2$ | Warfarin (N=6043) | 9484.6 | 45 | 47.5 | 1.0 (ref) | |
| | Rivaroxaban (N=5547) | 7760.2 | 26 | 33.5 | 0.68 (0.41–1.11) | 0.13 |
| eGFR ≤50 mL/min/1.73m$^2$ | Warfarin (N=1086) | 1538.1 | 18 | 117.0 | 1.0 (ref) | |
| | Rivaroxaban (N=889) | 1157.9 | 14 | 120.9 | 1.10 (0.53–2.29) | 0.79 |

**Note**: Method B was based on the Aberdeen algorithm. Severe AKI was defined as stage 2 or 3 AKI.

*Adjusted for age, sex, baseline eGFR (both as categorical and continuous variables), number of previous eGFR measurements at baseline, Townsend index, polymedication, smoking, body mass index, use of health services in the year before baseline (PCP visits, referrals and hospitalizations), comorbidity (ischemic heart disease, cancer, diabetes, heart failure and liver disease), frailty, $CHA_2DS_2VASc$ score, and use of antiplatelets/anti-infectives.

AKI, acute kidney injury; CI, confidence interval; eGFR, estimated glomerular filtration rate; HR, hazard ratio; PCP; primary care practitioner

**Supplementary Table 6**. Incidence rate per 10,000 person-years of severe AKI (using AKI Method B – see footnote) and HR (95% CI) comparing AKI in the rivaroxaban vs. warfarin cohorts, overall and stratified by baseline renal function (as-treated analysis).

| Baseline eGFR | Cohort | Person-years | Incident severe AKI cases (N) | Incidence rate per 10,000 person-years | HR* (95% CI) | *p* value |
|---|---|---|---|---|---|---|
| Any eGFR | Warfarin (N=7225) | 14,580.6 | 76 | 52.12 | 1.0 (ref) | |
| | Rivaroxaban (N=7020) | 11,861.3 | 49 | 41.31 | 0.82 (0.57–1.18) | 0.29 |
| eGFR >50mL/min/1.73m$^2$ | Warfarin (N=6119) | 12,563.2 | 52 | 41.39 | 1.0 (ref) | |
| | Rivaroxaban (N=6057) | 10,390.5 | 35 | 33.68 | 0.82 (0.53–1.27) | 0.38 |
| eGFR ≤50 mL/min/1.73m$^2$ | Warfarin (N=1106) | 2017.5 | 24 | 118.96 | 1.0 (ref) | |
| | Rivaroxaban (N=963) | 1470.9 | 14 | 95.18 | 0.85 (0.43–1.69) | 0.65 |

**Note**: Method B was based on the Aberdeen algorithm. Severe AKI was defined as stage 2 or 3 AKI.
*Adjusted for age, sex, baseline eGFR (both as categorical and continuous variables), number of previous eGFR measurements at baseline, Townsend index, polymedication, smoking, body mass index, use of health services in the year before baseline (PCP visits, referrals and hospitalizations), comorbidity (ischemic heart disease, cancer, diabetes, heart failure and liver disease), frailty, $CHA_2DS_2VASc$ score, and use of antiplatelets/anti-infectives.
AKI, acute kidney injury; CI, confidence interval; eGFR, estimated glomerular filtration rate; HR, hazard ratio; PCP; primary care practitioner

**Supplementary Table 7.** Incidence rate per 10,000 person-years of AKI and HR (95% CI) comparing AKI in the rivaroxaban vs. warfarin cohorts, stratified by baseline renal function (ITT analysis): Fine and Gray models.

| Cohort | Person-years | Incident AKI cases (N) | Incidence rate per 10,000 person-years | SHR* (95% CI) | *p* value |
|---|---|---|---|---|---|
| **AKI Method A** | | | | | |
| Warfarin (N=7129) | 20,027.6 | 136 | 67.9 | 1.00 (ref) | |
| Rivaroxaban (N=6436) | 13,917.5 | 113 | 81.2 | 1.15 (0.89–1.47) | 0.29 |
| **AKI Method B (any AKI)** | | | | | |
| Warfarin (N=7129) | 19,492.2 | 457 | 234.5 | 1.00 (ref) | |
| Rivaroxaban (N=6436) | 13,686.8 | 266 | 194.4 | 0.77 (0.66–0.89) | <0.01 |
| **AKI Method B (severe AKI)** | | | | | |
| Warfarin (N=7129) | 19,492.2 | 104 | 53.4 | 1.00 (ref) | |
| Rivaroxaban (N=6436) | 13,686.8 | 54 | 39.5 | 0.72 (0.51–1.00) | 0.05 |

**Note**: Method A was based on Read codes, and Method B was based on the Aberdeen algorithm. Severe AKI was defined as stage 2 or 3 AKI. (stage 1 was included as a competing risk).

*Adjusted for age, sex, baseline eGFR (both as categorical and continuous variables), number of previous eGFR measurements at baseline, Townsend index, polymedication, smoking, body mass index, use of health services in the year before baseline (PCP visits, referrals and hospitalizations), comorbidity (ischemic heart disease, cancer, diabetes, heart failure and liver disease), frailty, $CHA_2DS_2VASc$ score, and use of antiplatelets/anti-infectives.

AKI, acute kidney injury; CI, confidence interval; eGFR, estimated glomerular filtration rate; SHR, sub-distribution hazard ratio; PCP; primary care practitioner

**Supplementary Table 8**. Incidence rate per 10,000 person-years of AKI and HR (95% CI) comparing AKI in the rivaroxaban vs. warfarin cohorts, overall and stratified by baseline renal function: more stringent method B criteria.

| Baseline eGFR | Cohort | Person-years | Incident AKI cases (N) | Incidence rate per 10,000 person-years | HR* (95% CI) | *p* value |
|---|---|---|---|---|---|---|
| **AKI algorithm (meeting criterion 2 or 3)**† | | | | | | |
| Any eGFR | Warfarin (N=7129) | 20,134.2 | 60 | 29.8 | 1.0 (ref) | |
| | Rivaroxaban (N=6436) | 14,029.4 | 21 | 15.0 | 0.50 (0.30–0.84) | 0.01 |
| eGFR >50 | Warfarin (N=6043) | 17,455.3 | 38 | 21.8 | 1.0 (ref) | |
| | Rivaroxaban (N=5547) | 12,236.7 | 15 | 12.3 | 0.53 (0.28–1.00) | 0.05 |
| eGFR ≤50 | Warfarin (N=1086) | 2678.8 | 22 | 82.1 | 1.0 (ref) | |
| | Rivaroxaban (N=889) | 1792.7 | 6 | 33.5 | 0.36 (0.14–0.97) | 0.04 |
| **AKI algorithm (meeting criterion 3)**‡ | | | | | | |
| Any eGFR | Warfarin (N=7129) | 20,159.0 | 37 | 18.4 | 1.0 (ref) | |
| | Rivaroxaban (N=6436) | 14,041.6 | 11 | 7.8 | 0.41 (0.20–0.84) | 0.01 |
| eGFR >50 | Warfarin (N=6043) | 17,478.0 | 19 | 10.9 | 1.0 (ref) | |
| | Rivaroxaban (N=5547) | 12,243.7 | 9 | 7.3 | 0.70 (0.29–1.67) | 0.42 |
| eGFR ≤50 | Warfarin (N=1086) | 2681.1 | 18 | 67.1 | 1.0 (ref) | |
| | Rivaroxaban (N=889) | 1797.9 | 2 | 11.1 | 0.15 (0.03–0.71) | 0.02 |

**Note**: Method A was based on Read codes, and Method B was based on the Aberdeen algorithm.

*Adjusted for age, sex, baseline eGFR (both as categorical and continuous variables), number of previous eGFR measurements at baseline, Townsend index, polymedication, smoking, body mass index, use of health services in the year before baseline (PCP visits, referrals and hospitalizations), comorbidity (ischemic heart disease, cancer, diabetes, heart failure and liver disease), frailty, $CHA_2DS_2VASc$ score, and use of antiplatelets/anti-infectives.

†Serum creatinine ≥1.5 times higher than the lowest creatinine within 7 days (criterion 2) or serum creatinine >26 µmol/L higher than the lowest creatinine within 48 hours (criterion 3).

‡Serum creatinine >26 µmol/L higher than the lowest creatinine within 48 hours (criterion 3).

AKI, acute kidney injury; CI, confidence interval; eGFR, estimated glomerular filtration rate; HR, hazard ratio; PCP; primary care practitioner

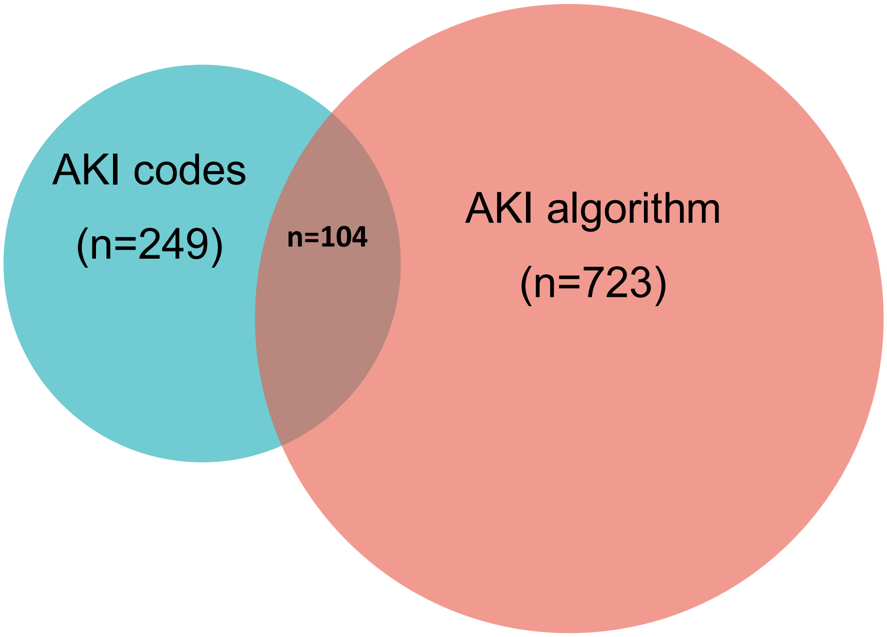


**Supplementary Figure 1.** AKI cases identified by the two case identification definitions.
AKI, acute kidney injury
